# Multimodal prediction of visual improvement in diabetic macular edema using real-world electronic health records and optical coherence tomography images

**DOI:** 10.64898/2026.04.23.26351616

**Authors:** Siqi Sun, Cindy X. Cai, Ruochong Fan, Saiyu You, Diep Tran, P. Kumar Rao, Marc A. Suchard, Yixin Wang, Cecilia S. Lee, Aaron Y. Lee, Linying Zhang

**Affiliations:** Institute for Informatics, Data Science and Biostatistics, Washington University in St. Louis, St. Louis, MO; Wilmer Eye Institute, Johns Hopkins School of Medicine, Baltimore, MD; Department of Biomedical Informatics and Data Science, Johns Hopkins School of Medicine, Baltimore, MD; John F. Hardesty MD, Department of Ophthalmology and Visual Sciences, Washington University in St. Louis, St. Louis, MO; Department of Biostatistics, University of California, Los Angeles, Los Angeles, CA; Department of Statistics, University of Michigan, Ann Arbor, Ann Arbor, MI; Department of Medicine, Washington University in St. Louis, St. Louis, MO

## Abstract

Multimodal learning has the potential to improve clinical prediction by integrating complementary data sources, but the incremental value of imaging beyond structured electronic health record (EHR) data remains unclear in real-world settings. We developed a multimodal survival modeling framework integrating optical coherence tomography (OCT) and EHR data to predict time to visual improvement in patients with diabetic macular edema (DME), and evaluated how different ophthalmic foundation model representations contribute to prognostic performance.

In a retrospective cohort of 973 patients (1,450 eyes) receiving anti-vascular endothelial growth factor therapy, we compared multimodal models combining 22,227 EHR variables with 196,402 OCT images, with OCT embeddings derived from three ophthalmic foundation models (RETFound, EyeCLIP, and VisionFM). The EHR-only model showed minimal prognostic discrimination (C-index 0.50 [95% CI, 0.45–0.55]). Incorporating OCT improved performance, with the magnitude of improvement depending on the representation. EHR+RETFound achieved the strongest performance (C-index 0.59 [0.54–0.65]), followed by EHR+EyeCLIP (0.57 [0.52–0.62]) and EHR+VisionFM (0.56 [0.51–0.61]). Multimodal models, particularly EHR+RETFound, demonstrated improved risk stratification with clearer separation of Kaplan–Meier curves.

Partial information decomposition revealed that prognostic information was dominated by modality-specific contributions, with OCT and EHR providing largely distinct signals and minimal shared information. The magnitude of OCT-specific contribution varied across foundation models and aligned with observed performance differences.

These findings indicate that OCT provides complementary prognostic value beyond structured clinical data, but gains are modest and depend strongly on representation choice. Our results highlight both the promise of multimodal modeling for personalized prognosis and the need for rigorous, context-specific evaluation of foundation models in real-world clinical settings.

## Introduction

Clinical decision-making is inherently multimodal. Clinicians must synthesize diverse sources of information, including demographic and clinical variables such as age, sex, comorbidities, medication history, laboratory test results that are typically stored as structured data in electronic health record (EHR) databases, as well as medical imaging data such as X-rays, magnetic resonance imaging (MRI), and pathology images, which collectively inform disease diagnosis, treatment planning, and health outcomes. Multimodal learning methods aim to integrate these complementary data modalities and therefore have the potential to improve prognostic accuracy and better support clinical decision-making^1–5^.

Multimodal learning is particularly promising in ophthalmology, where clinical decision-making heavily relies on imaging, such as fundus photography and optical coherence tomography (OCT)^6,7^. In this study, we focus on diabetic macular edema (DME), a leading cause of vision impairment among working-age adults worldwide^8^. OCT captures key morphological features of DME, including intraretinal and subretinal fluid accumulation, retinal thickening, and disruptions of retinal microstructure, which are critical for disease monitoring and assessment of treatment response^9,10^. Although anti-vascular endothelial growth factor (anti-VEGF) therapy is effective for treating DME, treatment response varies substantially across patients, making early prediction of visual improvement important for personalized risk stratification and treatment planning^11,12^.

Existing prognostic studies in DME have primarily relied on clinical and demographic variables, which do not capture retinal structural features visible on OCT^12–14^. Consequently, the incremental prognostic value of imaging beyond routinely available EHR data remains unclear^15^. Existing multimodal studies combining OCT and clinical variables for prognostic modeling have largely been conducted in curated research cohorts with limited sample sizes and standardized imaging protocols. These settings do not reflect real-world clinical practice, where patient populations are heterogeneous, follow-up duration is irregular, and imaging quality varies due to differences in devices and acquisition practices^16–18^. As a result, it remains an open question whether OCT provides meaningful and robust prognostic value beyond EHR in real-world settings.

Modeling multimodal clinical data presents additional challenges due to the high dimensionality of imaging features relative to sample size, which can lead to overfitting and poor generalizability. Recent advances in foundation models have enabled large-scale representation learning from medical images without task-specific feature engineering^19,20^. In clinical imaging, this promise is especially attractive because foundation models can reduce reliance on manual feature engineering and improve model generalizability across data sources^21,22^. Several ophthalmic foundation models have been developed to generate representations for various ophthalmic imaging modalities, including OCT. For example, RETFound is the first published ophthalmic foundation model trained on 1.6 million retinal images, including fundus photography and OCT, across 37,401 individuals from a single hospital in the United Kingdom^23^. VisionFM was subsequently released and pretrained on 3.4 million retinal images across 8 imaging modalities from 560,457 individuals internationally^24^.

EyeCLIP is a multimodal vision-language ophthalmic foundation model pretrained on 2.8 million ophthalmic images from 11 imaging modalities paired with partial clinical text to learn shared cross-modal representations^25^. While these models have demonstrated strong performance in imaging and cross-modal generation (e.g., generating clinical report from image) tasks, their clinical utility for real-world prognostic modeling, particularly in combination with structured EHR data, remains largely unexplored. To the best of our knowledge, no studies have systematically evaluated the incremental prognostic value of OCT in addition to structured EHRs for visual improvement prediction; nor have studies assessed whether the prognostic value of OCT representations differs across ophthalmic foundation models.

In parallel, most multimodal prediction studies focus on binary or continuous outcomes, whereas time-to-event (i.e., survival) modeling remain underutilized outside of oncology^26–29^ and critical care settings^30^. Survival analysis offers key advantages for EHR-based studies. EHR data are characterized by irregular follow-up and variable observation windows, and survival analysis naturally handles right censoring. In addition, survival analysis preserves clinically meaningful timing of events. For example, a patient achieving visual improvement at 6 months after treatment initiation is clinically distinct from one who improves at 12 months, and survival analysis retains this temporal information. These properties are particularly important in real-world ophthalmology data, where care continuity and follow-up intervals vary substantially across patients.

In this study, we develop a multimodal survival prediction framework integrating real-world OCT and EHR data to predict time to visual improvement in patients with DME. We systematically evaluate OCT representations derived from multiple ophthalmic foundation models and quantify their incremental prognostic value beyond EHR variables. Our study provides insights into the potential of integrating real-world multimodal data to improve prognostic modeling in ophthalmology.

## Results

### Cohort characteristics

We adopted a retrospective, intent-to-treat, new-user cohort design (Figure 1b). The study cohort included 973 patients with DME initiated with anti-VEGF therapy, contributing 1,450 eyes (196,402 OCT B-scans) that initiated anti-VEGF therapy during 2018-2024 (Supplementary Figure S1). Baseline patient characteristics were summarized in Table 1.

**Fig. 1.**
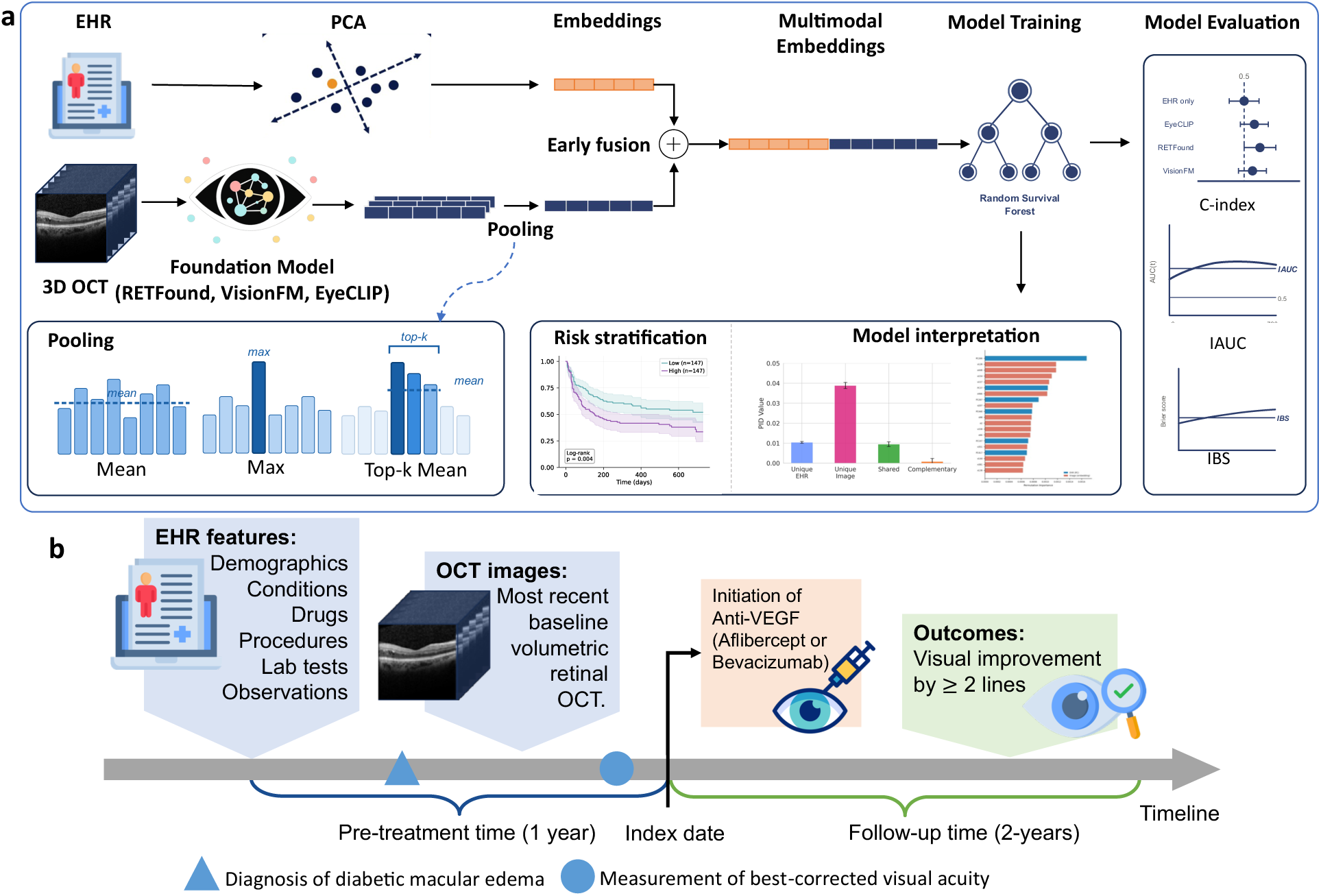
| Study overview. (a) Multimodal modeling framework. EHR features were compressed using principal component analysis (PCA) to generate EHR embeddings. OCT images were encoded using three pretrained ophthalmic foundation models: RETFound, VisionFM, and EyeCLIP. OCT embeddings were aggregated to eye-level representations using mean pooling, with max pooling and top-k mean pooling evaluated as sensitivity analyses. EHR and OCT embeddings were concatenated (i.e., early fusion) before fed into a random survival forest (RSF) model to predict visual improvement. Model performance was evaluated using Harrell’s C-index, integrated time-dependent AUC (IAUC), integrated Brier score (IBS). Downstream analyses include risk stratification and model interpretation. **(b) Study design and timeline.** The study cohort includes patients with DME receiving anti-VEGF therapy (aflibercept or bevacizumab) during the study period (2018-2024). Baseline data consist of 22,227 EHR-derived features and the most recent pre-treatment volumetric OCT. The index date is defined as the first injection of anti-VEGF therapy. The outcome is time to visual improvement (≥2 lines improvement in best-corrected visual acuity) over 2 years.

**Table 1.** Baseline Characteristics of the Study Cohort.

| Variable | Full cohort (973 patients / 1,450 eyes) |
| --- | --- |
| **Demographics (patient-level)** |  |
| Age (years) | 60.0 ± 11.8 |
| Female, n (%) | 512 (52.6%) |
| Race: White, n (%) | 455 (46.8%) |
| Race: Black or African American, n (%) | 464 (47.7%) |
| Race: Other or Unknown, n (%) | 54 (5.5%) |
| **Comorbidities (patient-level)** |  |
| Charlson Comorbidity Index | 5.5 ± 2.9 |
| Diabetes Comorbidity Severity (DCSI) | 5.3 ± 3.5 |
| CHADS2 score | 2.3 ± 1.4 |
| **Follow-up Period (patient-level)** |  |
| Follow-up time (days) | 274.3 ± 263.3 |
| **Ocular Characteristics (eye-level)** |  |
| Baseline VA (logMAR) | 0.5 ± 0.4 |
| Event (≥2-line VA improvement), n (%) | 718 (49.5%) |

Data are presented as mean ± SD or n (%). Demographics, comorbidities, and follow-up are reported at the patient level (n=973); ocular characteristics and event rate are reported at the eye level (n=1,450). VA: visual acuity; logMAR: logarithm of the minimum angle of resolution; CHADS2: Congestive heart failure, Hypertension, Age ≥ 75, Diabetes mellitus, Stroke/TIA; Other or Unknown race includes Asian (n=11), Other Pacific Islander (n=13), American Indian or Alaska Native (n=5), and race not reported in EHR (n=25).

At the patient level, the mean age was 60.0 years (SD, 11.8), and 52.6% of patients were female. The cohort was racially diverse, with 46.8% White, 47.7% Black or African American, and 5.5% other or unknown race. The mean Charlson Comorbidity Index was 5.5 (SD, 2.9), and the mean Diabetes Comorbidity Severity Index (DCSI) was 5.3 (SD, 3.5), indicating a substantial comorbidity burden. The mean CHADS2 score was 2.3 (SD, 1.4). The mean follow-up time was 274.3 days (SD, 263.3).

At the eye level, the mean baseline best-corrected visual acuity (BCVA) was 0.50 (SD, 0.40) logarithm of the minimum angle of resolution (logMAR). Most eyes (74.5%) received aflibercept injection. Overall, 718 of 1,450 eyes (49.5%) achieved a visual acuity improvement of at least two lines within 2 years of treatment initiation.

### Model performance

We compared three ophthalmic foundation models, three within-OCT pooling strategies (i.e., mean, max, and top-k mean) to aggregate image-level embeddings into eye-level embedding, and two cross-modality fusion strategies (early fusion, where modality-specific embeddings are concatenated prior to prediction model fitting, and late fusion, where modality-specific predictions are combined). Prognostic performance was evaluated using Harrell’s concordance index (C-index), integrated Brier score (IBS), and integrated time-dependent AUC (IAUC). All performance metrics were computed on the test set with bootstrapped confidence intervals (Figure 2).

**Fig. 2.**
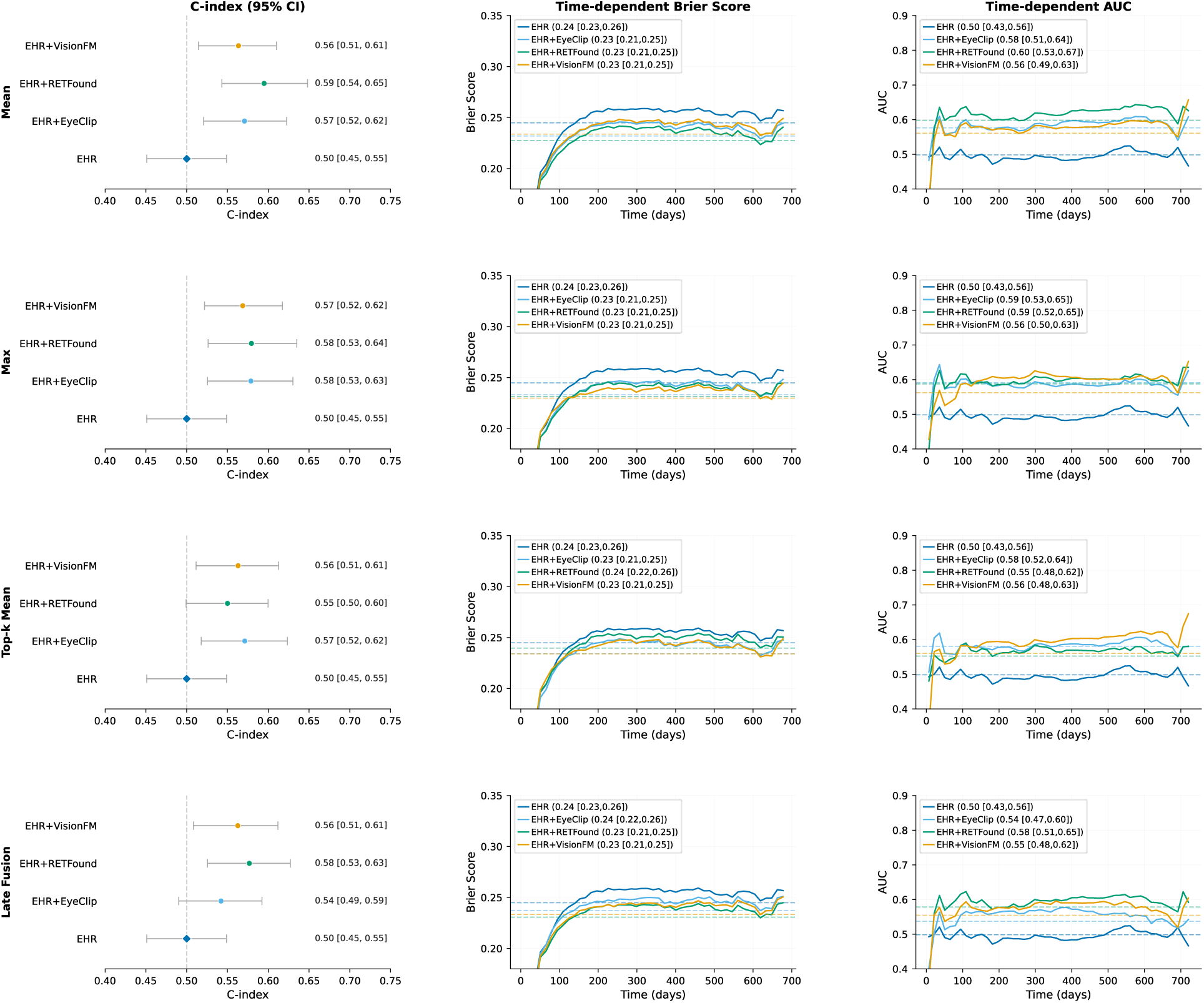
| Comparative performance of EHR-only and multimodal survival models across OCT pooling and modality fusion strategies. First three rows correspond to different OCT B-scan pooling strategies with early fusion: mean pooling (Mean), maximum pooling (Max), top-k mean pooling (Top-k Mean). The last row corresponds to late fusion with mean pooling. The left column shows Harrell’s C-index; higher values indicate better discrimination; the dashed vertical line at C-index=0.5 indicates random discrimination. The middle column shows time-dependent Brier score over the 730-day follow-up period; lower values indicate better calibration; dashed horizontal lines indicate the integrated Brier score (IBS) for each model. The right column shows time-dependent AUC over the 730-day follow-up period; higher values indicate better discrimination; dashed horizontal lines indicate the integrated AUC (IAUC) for each model.

Across experiments, performance was largely consistent across OCT pooling and modality fusion strategies, with no meaningful differences observed. Therefore, we present results from the early fusion with mean pooling configuration as the primary analysis. The EHR-only model showed minimal prognostic discrimination, with a C-index of 0.50 [0.45, 0.55], an IBS of 0.24 [0.23, 0.26], and an IAUC of 0.50 [0.43, 0.56], indicating performance close to random chance. Incorporating OCT improved performance, although the magnitude of improvement varied across ophthalmic foundation models. RETFound yielded the largest prognostic gains (C-index 0.59 [0.54, 0.65]; IBS 0.23 [0.21, 0.25]; IAUC 0.60 [0.53, 0.67]). EyeCLIP showed modest improvement over EHR-only model (C-index 0.57 [0.52, 0.62]; IBS 0.23 [0.21, 0.25]; IAUC 0.58 [0.51, 0.64]), while VisionFM demonstrated limited improvement (C-index 0.56 [0.51, 0.61]; IBS 0.23 [0.21, 0.25]; IAUC 0.56 [0.49, 0.63]).

To contextualize the added value of ophthalmic foundation models, we compared performance against a conventional pre-trained convolutional neural network baseline (ResNet50). Under the same analysis setting, EHR+ResNet50 achieved comparable performance to EyeCLIP and VisionFM but underperformed RETFound (Supplementary Table S3).

### Risk stratification by Kaplan-Meier analysis

To assess the clinical utility of model-derived risk scores, we stratified patients into high- and low-risk groups using the median risk score. Kaplan–Meier survival curves were constructed for patients in the test set (Figure 3).

**Fig. 3.**
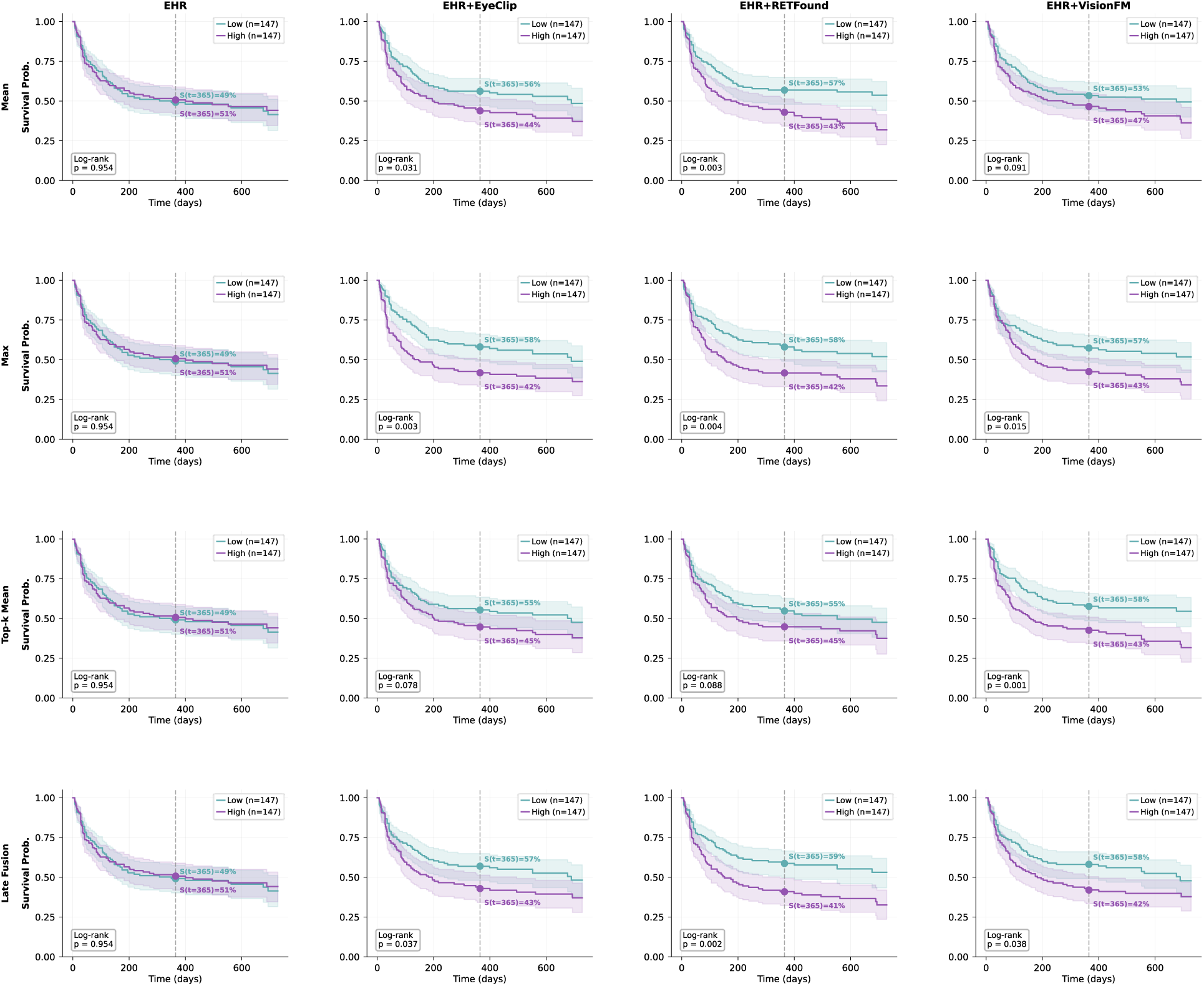
**| Risk stratification by multimodal models across OCT pooling and modality fusion strategies**. Kaplan–Meier survival curves for the test set stratified into low-risk and high-risk groups based on the median predicted risk score. First three rows correspond to different OCT B-scan pooling strategies with early fusion: mean pooling (Mean), maximum pooling (Max), top-k mean pooling (Top-k Mean). The last row corresponds to late fusion with mean pooling. Columns correspond to different models (from left to right): EHR-only, EHR+EyeCLIP, EHR+RETFound, and EHR+VisionFM. Shaded regions indicate 95% confidence intervals. Vertical dashed lines mark the 12-month timepoint, with corresponding survival probabilities annotated. Log-rank *p* values indicate the statistical significance of survival differences between risk groups.

Multimodal models have significantly improved risk stratification over EHR-only model across pooling and fusion strategies. Using the early fusion with mean pooling as an example, EHR+RETFound showed the strongest separation between low- and high-risk survival curves (log-rank test p = 0.003), with estimated 2-year probabilities of visual improvement 32% for the high-risk group and 54% for the low-risk group. EHR+EyeCLIP also showed significant separation, although weaker than RETFound (p = 0.031). In contrast, EHR-only and EHR+VisionFM showed limited separation (p = 0.954 and p = 0.091, respectively). The relative ranking of multimodal models in risk stratification was consistent with their performance metrics.

### Partial information decomposition (PID) of multimodal prognostic contributions

To characterize how OCT and EHR modalities contribute to prediction, we applied PID^31–33^ to partition modalities’ outcome-relevant information into four components: EHR-unique information, OCT-unique information, shared information captured by both modalities, and complementary information available only from their combination (Figure 4).

**Fig. 4.**
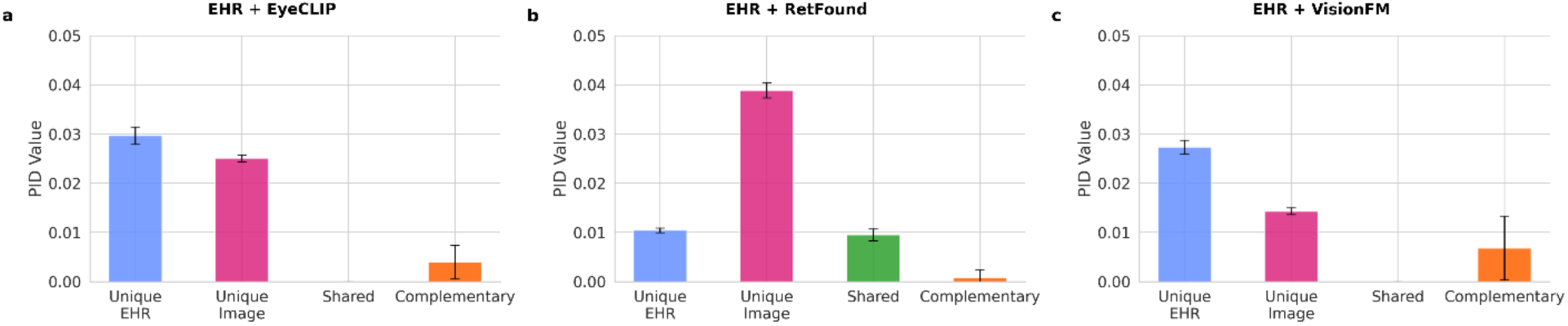
**| Partial information decomposition of prognostic information across modalities**. Bar plots show the decomposition of total prognostic information (measured by unique information, UUU, in bits) into modality-specific and joint components for multimodal survival models combining electronic health record (EHR) data with optical coherence tomography (OCT) embeddings derived from (a) EyeCLIP, (b) RETFound, and (c) VisionFM. Across models, OCT embeddings, particularly RETFound, contribute substantial unique prognostic information beyond EHR, while shared and complementary components remain relatively small.

Across all three ophthalmic foundation models, multimodal predictive information was dominated by unique contributions from individual modalities, with minimal shared information. This indicates that EHR and OCT features largely capture distinct aspects of patient health relevant to visual improvement, rather than overlapping signals.

In addition, complementary (synergistic) information was present but relatively small compared to unique components across all models, suggesting that while joint modeling provides some additional benefit, most predictive signal is captured independently by each modality.

The relative contributions of OCT-unique information varied substantially across foundation models. RETFound showed the largest OCT-unique contribution among the evaluated foundation models, exceeding the EHR-unique information. In contrast, EyeCLIP demonstrated more balanced contributions from both modalities, with smaller OCT-unique information compared to RETFound. VisionFM exhibited the lowest OCT-unique contribution, with EHR-unique information outweighing the OCT-unique information.

These PID patterns indicate that the incremental prognostic value of OCT is primarily driven by modality-specific information, and that the magnitude of this contribution is highly dependent on the choice of ophthalmic foundation model, with RETFound providing the most informative OCT representations.

### Feature importance and interpretation

To better understand the differences across multimodal models, we examined feature importance rankings in the multimodal models (Figure 5). The multimodal model with RETFound showed greater dominance of image embeddings among the highest-ranked variables. The other two multimodal models with EyeCLIP and VisionFM showed a more balanced distribution between EHR embeddings and image embeddings. These patterns were consistent with the PID analysis and overall model performance, suggesting that RETFound extracts more informative OCT representations that contribute substantially to prediction, whereas EyeCLIP and VisionFM rely more heavily on structured EHR features.

**Fig. 5.**
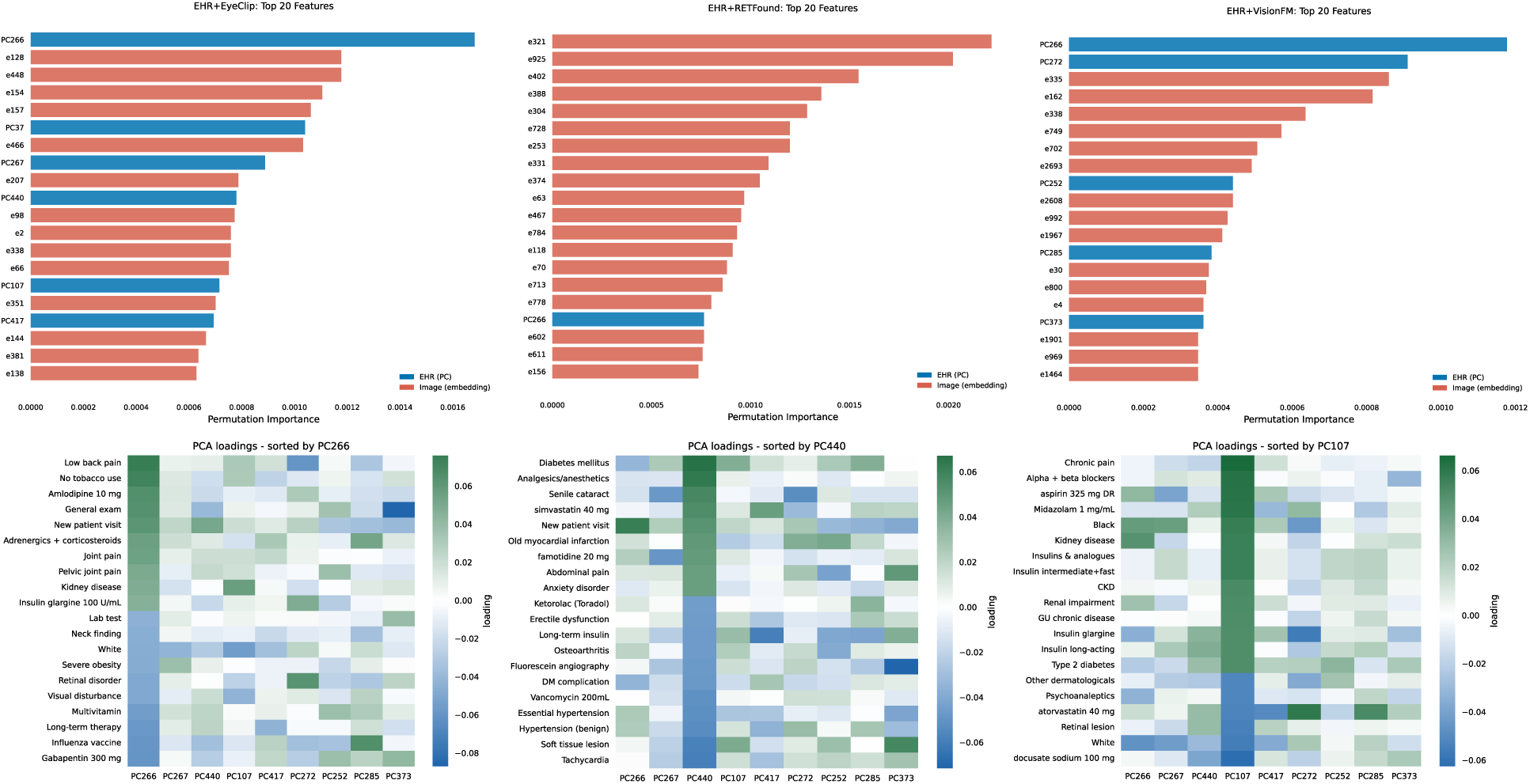
| Feature importance and interpretation. (a-c) Top 20 most important features across three multimodal models (a) EHR+EyeCLIP, (b) EHR+RETFound, and (c) EHR+VisionFM. Blue = EHR embeddings; Orange = OCT embeddings. (d-f): Interpretation of selected predictive EHR-derived principle. Each panel displays the top 20 variables by absolute loading for one principal component, sorted from highest to lowest loading in that PC, with all nine PCs shown as columns. Color intensity reflects PCA loading magnitude. Each PC captures distinct yet partially overlapping clinical themes: (d) PC266 reflects general outpatient care and chronic pain; (e) PC440 reflects cardiovascular and metabolic conditions; (f) PC107 reflects renal and diabetic conditions.

Inspection of the highest-loading raw variables within important EHR principal components suggested that these components captured clinically meaningful themes related to chronic diseases, such as diabetes, cardiovascular diseases, chronic pain, and renal diseases, as well as ocular disease severity, as indicated by fluorescein angiography and cataract (Figure 5d-f & Supplementary Figure S3).

## Discussion

This study developed a multimodal modeling framework integrating OCT imaging and EHR data to predict visual improvement among patients with diabetic macular edema in real-world clinical practice. We found that incorporating OCT representations improved prognostic performance over EHR-only models, with consistent gains across discrimination, calibration, and risk stratification metrics. Among the evaluated models, RETFound yielded the largest improvements, while EyeCLIP and VisionFM showed more modest gains. These improvements translated into clinically meaningful separation of Kaplan–Meier curves, suggesting potential utility for patient risk stratification in routine care.

A key finding of this study is that the incremental prognostic value of OCT is primarily driven by modality-specific (unique) information rather than shared information with EHR variables. Partial information decomposition demonstrated that EHR and OCT contributed largely distinct, non-overlapping signals, with minimal shared information across all models. This suggests that imaging captures aspects of disease biology and retinal structure that are not well represented in structured clinical data. While complementary (synergistic) information between modalities was present, it was relatively small compared to the unique contributions, indicating that most predictive signal is derived independently from each modality rather than from their interaction. These findings refine the understanding of multimodal learning in clinical ophthalmology settings, highlighting that performance gains arise from aggregating distinct sources of information rather than exploiting their interactions.

The magnitude of OCT-derived prognostic value varied substantially across ophthalmic foundation models. RETFound demonstrated the largest OCT-unique contribution and strongest overall performance, consistent with feature importance analyses showing dominance of imaging embeddings in the multimodal model. In contrast, EyeCLIP exhibited more balanced contributions between modalities, and VisionFM showed relatively limited OCT-specific signal. These differences likely reflect variation in pretraining strategies and data composition. RETFound’s self-supervised training on two retinal imaging modalities, one of which is OCT, may better capture fine-grained structural features in OCT that are relevant to DME, whereas EyeCLIP’s vision-language alignment and VisionFM’s multi-modality pretraining may prioritize broader semantic or cross-modal representations at the expense of OCT-specific prognostic features^23–25^. These observations suggest that the clinical utility of foundation models may depend on how well the pretraining strategy and training data composition align with the downstream clinical task.

Our analysis also highlights limitations of current OCT representation strategies. In this study, OCT volumes were represented by aggregating embeddings from individual B-scans using pooling operations. While performance was robust across pooling strategies, this approach treats volumetric data as unordered collections of slices and may discard spatial relationships across the retina. Given that OCT images are inherently volumetric^9^, and clinically meaningful features, such as retinal fluid distribution and retinal layer deformation, often span multiple B-scans, future work leveraging volumetric OCT foundation models may better capture these structural dependencies and further improve prognostic performance^34,35^.

Despite consistent improvements from multimodal integration, the overall prediction performance remained modest. This likely reflects the complexity and heterogeneity of real-world clinical data. For example, OCT scans are acquired at variable times relative to treatment initiation, and imaging protocols differ across devices in resolution and number of B-scans per session. These sources of variability can weaken the alignment between imaging and clinical data, making it more difficult for models to extract complementary signals across modalities^36^. In addition, treatment adherence in routine care is highly variable (Supplementary Figure S2). While many patients persist with their initial therapy, a substantial proportion experience delays, switch treatments, or discontinue early, leading to irregular injection intervals and attrition over time. Such real-world treatment patterns can introduce exposure misclassification, complicating the estimation of treatment-specific prognostic signals. Furthermore, inconsistent follow-up is common in real-world settings; missed or delayed visits may result in incomplete or delayed capture of visual acuity outcomes, further degrading model performance.

Beyond ophthalmology, these findings have broader implications for the development of multimodal clinical artificial intelligence development. The growing availability of pre-trained foundation models across data modalities has lowered the barriers to multimodal learning, and evidence suggesting that learning from multimodal data leads to better performance has exploded in the past several years^21^. However, these reported gains may be influenced by publication bias, and accumulating evidence indicates that the benefits of multimodal learning and foundation models vary across clinical domains and prediction tasks. For example, a recent multimodal study in cardiovascular disease detection found that adding chest radiographs to electrocardiogram data produced little improvement in prognostic performance, with most of the prognostic information shared across modalities rather than uniquely contributed by imaging^37^. In addition, foundation models have, in some settings, demonstrated lower generalizability compared to locally trained models^38^. These findings, along with our results, underscore the persistent need for rigorous and context-specific evaluation of the clinical utility of generalist foundation models and multimodal learning in real-world settings.

This study has several limitations. First, the cohort was derived from a single health system in the Midwestern United States, and external validation across health systems, imaging devices, and clinical practice settings will be necessary to assess transportability. Second, the analysis is retrospective, and prospective evaluation, especially randomized comparison against human clinicians, is needed to confirm the model’s clinical utility^39^. Third, although we focused on leveraging representations from pretrained foundation models without task-specific fine-tuning or domain adaptation, which may further improve downstream model performance^40^. Finally, our interpretation analyses were conducted at the representation level between modalities and did not localize specific anatomical features within OCT images that drive prognostic performance^41,42^.

In summary, we demonstrate that OCT imaging provides distinct and complementary prognostic information beyond structured EHR data for predicting visual improvement in diabetic macular edema, with the magnitude of benefit strongly dependent on the choice of foundation model. While the overall performance gains are modest in real-world settings, these findings highlight both the promise of multimodal integration and the importance of understanding how different data modalities contribute to clinical prediction.

## Methods

### Data sources

Data were obtained from the Washington University/BJC HealthCare (WashU) EHR data warehouse. WashU EHR has over <u>5.7 million patients</u> for over <u>25 years</u> across 14 hospitals within BJC HealthCare, a regional health system focused on delivering health services to residents primarily in the greater St. Louis, southern Illinois and mid-Missouri regions. WashU EHR database includes demographics, conditions, medications, laboratory results, procedures, and observations, standardized to the Observational Medical Outcomes Partnership Common Data Model (OMOP CDM) v5.4. OCT images were extracted from the clinical ophthalmic imaging database, accessed through Continuum. OCT images were acquired by either Cirrus (Carl Zeiss Meditec) or Spectralis (Heidelberg Engineering) devices. This study was approved by the Institutional Review Board (IRB) under IRB protocol 202502158.

### Cohort definition

The study cohort was defined with the following inclusion criteria (Figure 1b and Supplementary Figure S1): (1) study period between January 1, 2018 and December 31, 2024; (2) diagnosis of diabetic macular edema (DME) during the study period; (3) receipt of intravitreal anti-VEGF injection (aflibercept or bevacizumab); and (4) availability of injection laterality (OD/OS/OU) to identify the treated eye, with OU entries duplicated and split into OD and OS to create eye-level observations. Exclusion criteria were: (1) missing baseline visual acuity (VA), defined as no VA measurement within 365 days prior to the first injection or non-convertible VA values; (2) missing OCT imaging, defined as no baseline OCT scans within 365 days prior to the first injection. The index date was defined the date of the first anti-VEGF injection. All VA measurements were converted to logarithm of the minimum angle of resolution (logMAR) units for analysis.

The outcome was defined as the days from the first injection to the first post-treatment VA measurement demonstrating a ≥2-line improvement in VA relative to baseline within 730 days of follow-up. Eyes not reaching the endpoint were censored at the day with the last available VA measurement or 730 days, whichever comes first. The medical codes used to define clinical concepts were included in Supplementary Table S1 and Supplementary Table S2.

### OCT embeddings and pooling strategies

Each B-scan is resized to 224×224 pixels and embedded using pretrained ophthalmic foundation models: RETFound (1024-dimensional), VisionFM (3072-dimensional), and EyeCLIP (512-dimensional). Embeddings are computed per B-scan and then aggregated using mean pooling, max pooling, or top-k mean pooling to obtain eye-level OCT embedding. The number of B-scans per eye depended on the devices (25 for Spectralis; 128 for Cirrus). Additional details about OCT data preprocessing, embedding extraction, pooling strategies are provided in Supplementary Note S1.

### EHR embeddings with principal component analysis (PCA)

Electronic health records were extracted from the OMOP Common Data Model and included structured clinical covariates from routine care. After excluding identifiers, outcome variables, treatment indicators, and other non-predictor fields, principal component analysis (PCA) was then applied to the remaining EHR features. The minimum number of principal components (PC) explaining 95% of the cumulative variance in the training data was retained, which included 445 PCs, and used as the EHR input to the survival model. All preprocessing steps were fit on the training set only and then applied to the test set. PCA was fitted within the training folds and applied to test fold in a 5-fold cross-validation to prevent information leakage^43,44^.

### Model development and evaluation

All comparisons used a patient-level train/test split (80% train, 20% test) to prevent leakage between eyes of the same patient^45–47^. We used a random survival forest (RSF)^48^ for all experiments because it is a non-parametric survival model that can accommodate non-linear effects and interactions in high-dimensional covariates without requiring proportional hazards assumptions. RSF hyperparameters (n_estimators, min_samples_leaf, max_features) were selected via five-fold cross-validation on the training set using grid search; the best-performing configuration was then refit on the full training set. The evaluation metrics included Harrell’s C-index^49^, integrated time-dependent AUC (IAUC)^50^, and integrated Brier score (IBS)^51^. Bootstrap resampling of the test set (B=1000) was used to estimate 95% confidence intervals for all evaluation metrics. Risk groups were defined by median predicted risk score, and survival differences between groups were assessed using the log-rank test^52^.

### Multimodal data fusion strategies

For fusion sensitivity, we compared early fusion (*EF*) versus late fusion (*LF*). In early fusion, we concatenated EHR features *x_i_* ∈ *R^dx^* and OCT features *Z_i_* ∈ *R^dz^* into a single input *U_i_* = [*X_i_*; *Z_i_*] and trained one RSF to output a risk score *r*^EF^, evaluated by test-set Harrell’s C-index *C*_EF_.

In late fusion^53^, we trained two separate RSF models to produce modality-specific risk scores 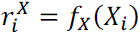 and 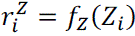, computed validation C-indices 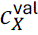 and 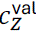, converted them to nonnegative weights 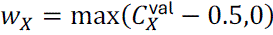 and 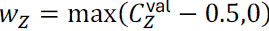, and formed the fused test risk score

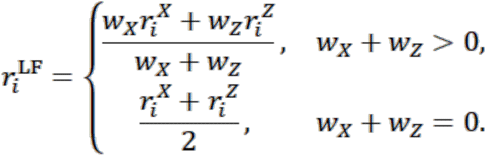

We then evaluated the fused model using test-set Harrell’s C-index *C_LF_*. Across all sensitivity settings, we compared the ordering of OCT representations and the stability of incremental value conclusions.

### Partial information decomposition (PID)

PID provides a principled framework for quantifying how multiple input modalities jointly contribute to predicting an outcome by decomposing total information into shared (redundant), unique, and complementary (synergistic) components^31–33^. We adopted the scalable estimation approach proposed by Liang et al.^33^, which enable PID computation for high-dimensional multimodal data using neural parameterization and marginal-matching constraints. The implementation was adopted from Choi et al.^37^, and was briefly re-iterated as follows.

Let *X*denote the EHR embedding from PCA, *Z*denote the eye-level OCT embedding, and *Y* ∈ {0,1} denote the event indicator. We first trained three probabilistic multilayer perceptron (MLP) classifiers to estimate

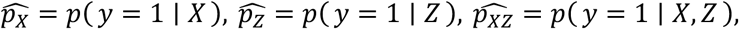

each optimized with binary cross-entropy loss. We then trained a *Q*-estimator (a two-branch MLP) to learn a constrained joint distribution *q*(*X*, *Z*, *Y*) subject to marginal constraints implied by 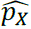 and 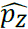 using Sinkhorn–Knopp updates, and the model parameters were optimized to minimize the mutual information objective

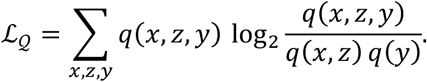

On the held-out test set, we computed the PID components {EHR-unique, OCT-unique, shared, complementary}, corresponding to information uniquely provided by the EHRs, uniquely provided by OCT, redundant information shared by both modalities, and synergistic information available only from the joint representation. We repeated the computation five times with different random permutations of the test set and report mean±SD across runs.

### Feature importance with permutation

To quantify the relative contribution of each feature to model performance, we assessed feature importance by applying the final trained model on the held-out test set, where each feature was randomly permuted across samples while holding other features unchanged, and the resulting change in model performance was computed and interpreted as the feature importance score. This procedure was repeated five times per feature, and the mean permutation importance across repetitions was used as the feature-level importance score.

### Statistical analysis

Continuous variables are presented as mean ± standard deviation or median [interquartile range], as appropriate, and categorical variables are presented as n (%). Model performance was assessed on the held-out test set, and 95% confidence intervals were obtained using nonparametric bootstrap resampling with 1,000 replicates. For risk stratification analyses, Kaplan–Meier curves were estimated for groups defined by the median predicted risk score and compared using the log-rank test. All analyses were conducted in Python using pandas, NumPy, scikit-learn, scikit-survival, lifelines, and PyTorch.

### Ethical approval

The study was conducted in accordance with the Declaration of Helsinki and was approved by the Institutional Review Board of Washington University School of Medicine in St. Louis (IRB No. 202502158). A waiver of informed consent was granted because of the retrospective design and use of de-identified data.

## Data availability

The datasets generated and analyzed during this study are available from the corresponding author upon reasonable request, subject to institutional data sharing agreements.

## Acknowledgements

L.Z. was partially supported by internal funds from the Washington University Transdisciplinary Institute in Applied Data Sciences (TRIADS) Seed Grant Program (PJ000030883) and the Washington University Here and Next Research Grant (PJ000030799). Y.W. was supported in part by funding from the Office of Naval Research under grant N00014-23-1-2590, the National Science Foundation under grant No. 2310831, No. 2428059, No. 2435696, No. 2440954, a Michigan Institute for Data Science Propelling Original Data Science (PODS) grant, Two Sigma Investments LP, and LG Management Development Institute AI Research. C.S.L. was partly supported by NIH/NIA 2R01AG060942 and NIH OT2OD32644. A.Y.L. was partly supported by NIH OT2OD032644, NIH/NIA R01AG060942 and NIH/NIA U19AG066567, and Research to Prevent Blindness. The authors would like to acknowledge Shinji Naka from the Informatics Core Services (ICS) at Washington University for his guidance on querying EHR data.

## Author information

### Authors and Affiliations

**Institute for Informatics, Data Science and Biostatistics, Washington University in St. Louis, St. Louis, MO, USA**

Siqi Sun, Ruochong Fan, Saiyu You & Linying Zhang

**Wilmer Eye Institute, Johns Hopkins School of Medicine, Baltimore, MD, USA**

Cindy X. Cai & Diep Tran

**Department of Biomedical Informatics and Data Science, Johns Hopkins School of Medicine, Baltimore, MD, USA**

Cindy X. Cai

**Department of Ophthalmology and Visual Sciences, Washington University in St. Louis, St. Louis, MO, USA**

P. Kumar Rao, Cecilia S. Lee & Aaron Y. Lee

**Department of Biostatistics, University of California, Los Angeles, CA, USA**

Marc A. Suchard

**Department of Statistics, University of Michigan, Ann Arbor, MI, USA**

Yixin Wang

### Contributions

S.S. contributed to conceptualization, methodology, analysis and interpretation of results, and manuscript writing. C.X.C. contributed to data curation of visual acuity data and manuscript review and editing. R.F. contributed to data extraction and manuscript editing.

S.Y. contributed to OCT image extraction and data curation. D.T. contributed to data curation of visual acuity data. P.K.R., C.S.L., and A.Y.L. contributed to clinical interpretation and manuscript review. M.A.S. and Y.W. contributed to methodology and manuscript review and editing. L.Z. contributed to conceptualization, methodology, supervision, and manuscript review and editing. All authors reviewed and approved the final manuscript.

### Corresponding author

Correspondence to Linying Zhang

