## Supplementary Materials for "Multimodal prediction of visual improvement in diabetic macular edema using real-world electronic health records and optical coherence tomography images"

**Supplementary Figure S1. Cohort selection flowchart.**

Patients were identified from the WashU EHR database and sequentially screened based on study eligibility criteria. The final analytic cohort included 973 patients (1,450 eyes). NLP = no light perception; NI = no improvement; NPL = no perception of light; OCT = optical coherence tomography; EHR = electronic health record.

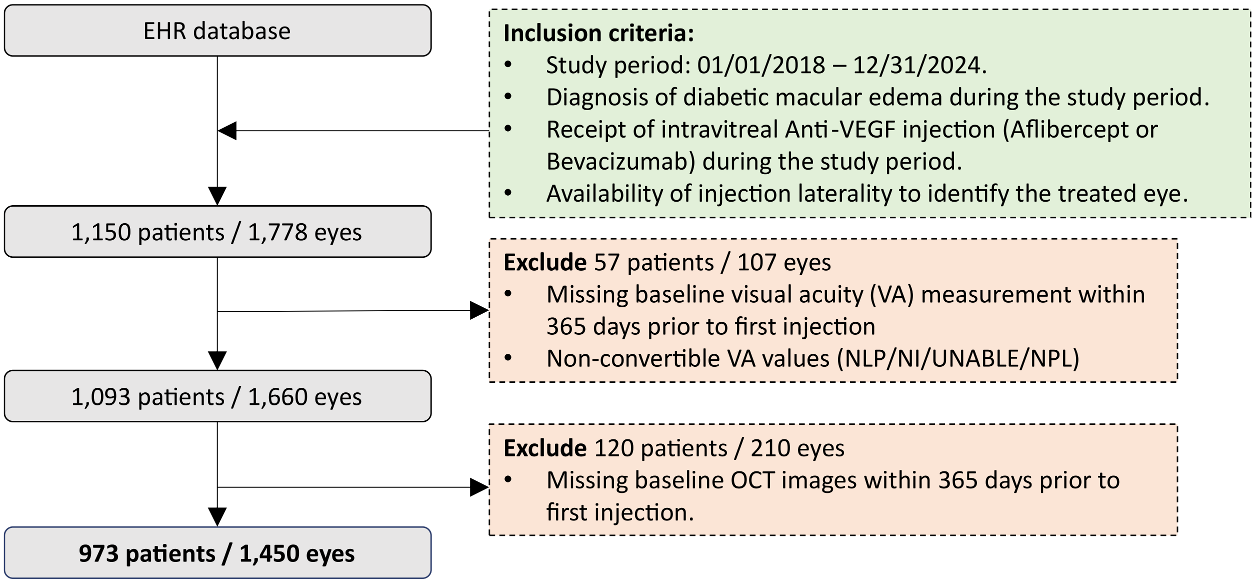


**Supplementary Table S1. Codes from standardized OMOP vocabularies used to define clinical concepts.**

| Clinical Concept | Concept ID | Concept Name | Source Code | Source Vocabulary | Concept Class Id |
|---|---|---|---|---|---|
| aflibercept | 40244266 | aflibercept | 1232150 | RxNorm | Ingredient |
| bevacizumab | 1397141 | bevacizumab | 253337 | RxNorm | Ingredient |
| ranibizumab | 19080982 | ranibizumab | 595060 | RxNorm | Ingredient |
| diabetic macular edema | 380097 | Macular edema due to diabetes mellitus | 312912001 | SNOMED | Disorder |
| Intravitreal injection | 2111025 | Intravitreal injection of a pharmacologic agent (separate procedure) | 67028 | | CPT4 |

**Supplementary Table S2. Medication IDs used to define anti-VEGF therapy from Epic Clarity database.**

| Medication Id | Medication Name |
|---|---|
| 112992 | Aflibercept 2 mg/0.05 ml intravitreal solution for injection |
| 205711 | Aflibercept 2 mg/0.05 ml intravitreal syringe |
| 223672 | Aflibercept 8 mg/0.07 ml intravitreal solution for injection |
| 108065 | Bevacizumab 100mg/4ml (25 mg/ml) intravenous solution |
| 186324 | Bevacizumab 2.5 mg/0.1 ml intravitreal syringe |
| 190037 | Bevacizumab 1.25 mg/0.05 ml intravitreal syringe |
| 204125 | Bevacizumab 3.25 mg/0.13 ml intravitreal syringe |
| 225658 | Bevacizumab 2.75 mg/0.11 ml intravitreal syringe |
| 503665 | Bevacizumab 25 mg/ml intravenous solution for intraocular use |
| 76790 | Ranibizumab 0.5 mg/0.05 ml intravitreal solution for injection |
| 117197 | Ranibizumab 0.3 mg/0.05 ml intravitreal solution for injection |
| 186862 | Ranibizumab 0.5 mg/0.05 ml intravitreal syringe |
| 195995 | Ranibizumab 0.3 mg/0.05 ml intravitreal syringe |

**Supplementary Figure S2. Real-world treatment adherence and transition patterns. (a)** the Sankey diagram shows patient flows between treatments at each injection. Aflibercept was the most commonly used agent at initiation (801 patients), followed by bevacizumab (363) and ranibizumab (69). At the second injection, most patients continued with the same therapy (e.g., 668 remained on aflibercept and 244 on bevacizumab), although a subset switched between agents or discontinued treatment (263 patients had no second injection). By the third injection, persistence remained highest for aflibercept (550 patients) and bevacizumab (180), with fewer patients continuing ranibizumab (47), while a substantial proportion (456 patients) did not receive a third injection. Overall, treatment patterns indicate moderate persistence with initial therapy, particularly for aflibercept, alongside notable attrition between injections. **(b)** Distribution of time gaps between consecutive injections. The majority of intervals between the first and second injections were relatively short, with a strong peak within approximately 2-3 months, although a long tail extending beyond several hundred days indicates variability in time gap between injections. A similar pattern was observed for the interval between the second and third injections.

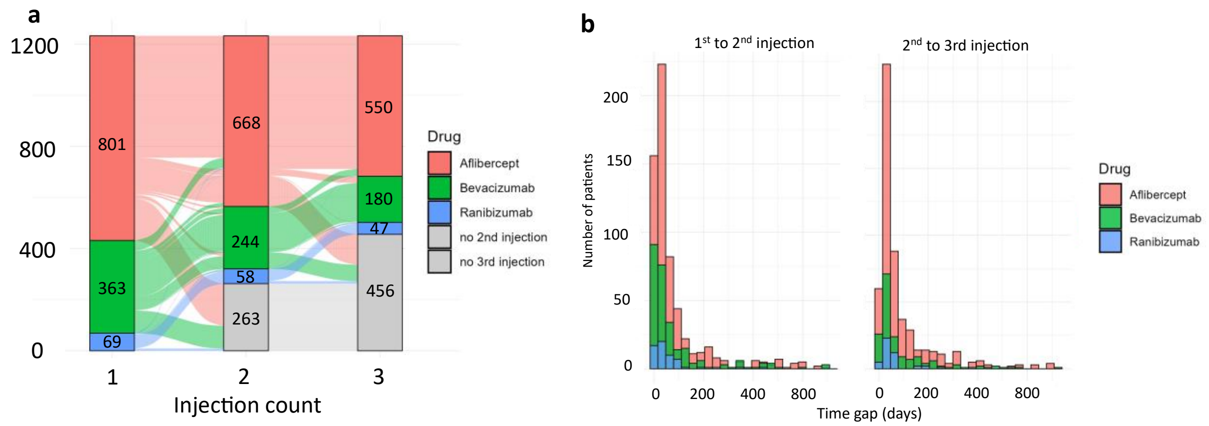


### Supplementary Note S1. Details of OCT embeddings

RETFound, VisionFM, and EyeCLIP were evaluated as frozen feature extractors for generating OCT embeddings, with key architectural characteristics summarized in the table below. The models differ in backbone architecture, pretraining objective, training data composition, and parameter scale.

Before embedding extraction, all OCT B-scan images were converted to RGB format, resized to 224 × 224 pixels and normalized using the model-specific input statistics. Each B-scan was then passed through the corresponding frozen visual encoder to obtain a single embedding vector.

To derive eye-level representations, embeddings were aggregated across all B-scans from the same eye using three strategies: mean pooling, max pooling and top-k mean pooling, which are described in details as follows.

For an eye $e$, let $Z_{e,s} \in R^d$ denote the embedding from B-scan$s \in \{1, \dots, n_e\}$, where $d$ is the embedding dimension and $n_e$ is the number of B-scans available for that eye. Because outcomes and covariates are defined at eye level, we constructed an eye-level embedding $Z_e \in R^d$ by aggregating $\{Z_{e,s}\}_{s=1}^{n_e}$ using three pooling strategies[1,2]:

(1) Mean pooling:

$$Z_e = \frac{1}{n_e} \sum_{s=1}^{n_e} z_{e,s}.$$

(2) Max pooling (elementwise): for feature dimension$j \in \{1, \dots, d\}$,

$$(Z_e)_j = \max_{s \in \{1, \dots, n_e\}} \left(z_{e,s}\right)_j.$$

(3) Top-$k$ mean pooling: we ranked B-scans by the $\mathrm{l}_2$ norm $\left\|z_{e,s}\right\|_2$, selected the $k_e = \min(k, n_e)$ scans with the largest norms, and computed

$$Z_e = \frac{1}{k_e} \sum_{s \in S_e^{(k_e)}} z_{e,s}.$$

where $S_e^{(k_e)}$ denotes the index set of the selected scans. In this study, we selected the top 5 B-scans in top-k pooling (K=5).

Mean pooling and max pooling are widely used, non-parametric and permutation-invariant aggregators for summarizing multiple instance (e.g., multi-slice or multi-scan) representations into a single sample-level embedding, and they are commonly adopted as standard baselines in multiple instance learning (MIL)[1] and set-structured modeling. Top-$k$ mean pooling is a related variant that first selects the $k$ highest-scoring instances and then averages their embeddings and has been used as an alternative MIL pooling operator when the prognostic signal is expected to concentrate in a subset of instances.

| | RETFound | VisionFM | EyeCLIP |
|---|---|---|---|
| Backbone | ViT-Large (ViT-L/16) | ViT-Base/(ViT-B/16) | ViT-Base/(ViT-B/32) |
| Pretraining method | MAE | iBOT | CLIP + MAE |
| Pretraining data | 736,442 OCT B-scans | 1,420,357 OCT B-scans | 54,126 OCT B-scans |
| Paradigm | Unimodal (OCT only) | Multimodal (modality-specific encoders) | Multimodal (unified encoder, vision-language) |
| Parameters | 329.5M | 96.3M | 87.8M |
| Output dimension | 1024 | 3072 | 512 |

MAE: Masked Autoencoder; iBOT: Image BERT pre-training with Online Tokenizer; CLIP: Contrastive Language–Image Pre-training.

**Supplementary Figure S3. Heatmap of PCA loadings for additional six important EHR-derived principal components.** Each panel displays the top 20 variables by absolute loading for one principal component, sorted from highest to lowest loading in that PC, with all nine PCs shown as columns. Color intensity reflects loading magnitude, with green indicating positive loadings and blue indicating negative loadings. Each PC captures distinct yet partially overlapping clinical themes: PC267 captures preventive care and metabolic management; PC417 captures acute procedural and cardiovascular events; PC272 reflects retinal and ophthalmic disease; PC252 captures diabetes pharmacotherapy; PC285 reflects chronic disease management and lifestyle factors; and PC373 highlights ophthalmic procedures and acute care settings.

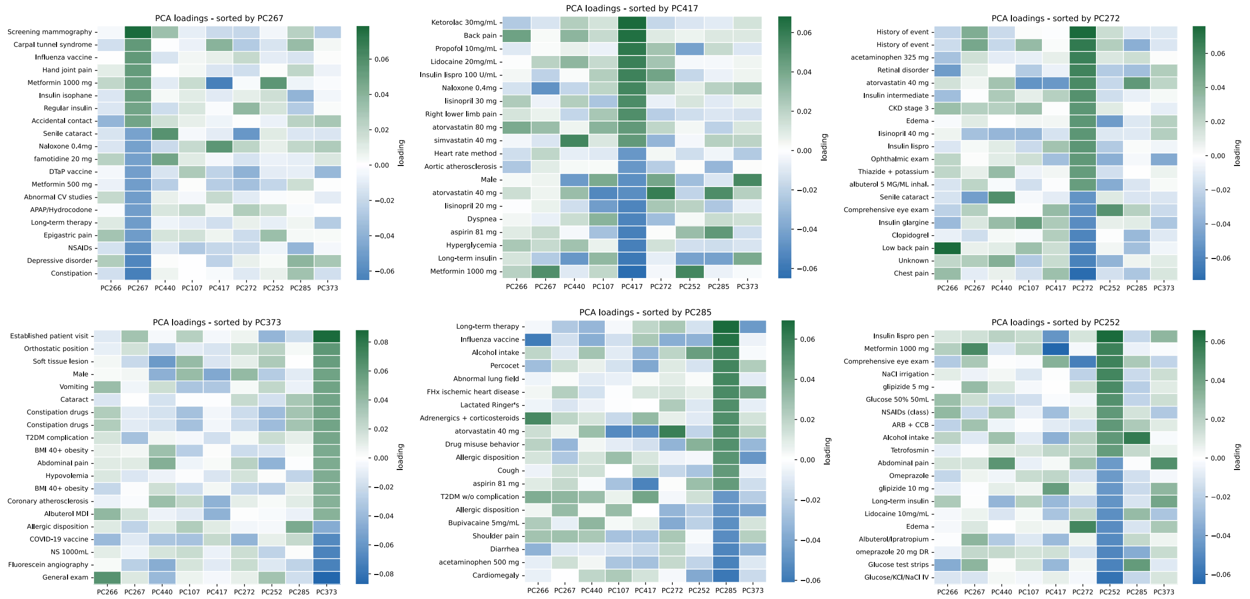


**Supplementary Table S3. Performance comparison between ophthalmic foundation models and a generalist imaging model for OCT representation**

To contextualize the added value of ophthalmic foundation models, we included a conventional computer vision baseline based on a standard convolutional neural network. This comparison was intended to distinguish whether the observed gains in multimodal performance arise from domain-specific pretraining (foundation models) or can be achieved using generic imaging models.

To do so, we applied ResNet50 initialized with ImageNet weights for OCT embedding generation. ResNet50[3] was used as a feature extractor without fine-tuning or survival supervision to provide a comparison with the ophthalmic foundation models used in the main analysis. Each OCT B-scan was resized to 224 × 224 pixels, converted to three-channel input, and normalized using the standard ImageNet preprocessing pipeline. The final classification layer was removed, and a 2048-dimensional feature vector was extracted from each B-scan. Eye-level OCT representations were then constructed by mean pooling across all available B-scans within the same eye. These ResNet50 eye-level embeddings were concatenated with EHR features using the same early-fusion pipeline as in the primary analysis and evaluated with the same random survival forest framework and test-set metrics were reported.

Under the same primary analysis setting (mean pooling and early fusion), the ResNet50 baseline achieved a C-index of 0.56 [0.51, 0.61], an IBS of 0.24 [0.22, 0.26], and an IAUC of 0.57 [0.50, 0.63]. Compared with the ophthalmic foundation models, EHR+ResNet50 showed lower prognostic performance than EHR+RETFound, while performing comparably to EHR+VisionFM and EHR+EyeCLIP. These results suggest that domain-specific pretraining may provide additional incremental prognostic value to OCT representations; however, the observed improvement is not statistically significant.

| Model | C-index | IBS | IAUC |
|---|---|---|---|
| EHR-only | 0.50 [0.45, 0.55] | 0.24 [0.23, 0.26] | 0.50 [0.43, 0.56] |
| EHR + ResNet50 | 0.56 [0.51, 0.61] | 0.24 [0.22, 0.26] | 0.57 [0.50, 0.63] |
| EHR + RETFound | 0.59 [0.54, 0.65] | 0.23 [0.21, 0.25] | 0.60 [0.53, 0.67] |
| EHR + EyeCLIP | 0.57 [0.52, 0.62] | 0.23 [0.21, 0.25] | 0.58 [0.51, 0.64] |
| EHR + VisionFM | 0.56 [0.51, 0.61] | 0.23 [0.21, 0.25] | 0.56 [0.49, 0.63] |
